# Changes in High-Sensitivity Troponin I Dynamics Pre and Post Parturition are Phenotype Specific

**DOI:** 10.1101/2025.10.21.25338264

**Authors:** Yaxin Li, He S. Yang, Zhengming Chen, Isla Racine, Damien Gruson, Allan S. Jaffe, Zhen Zhao

## Abstract

**Background:** Hypertensive disorders of pregnancy (HDP), including preeclampsia (PEC) and chronic/gestational hypertension (HTN), are major causes of maternal morbidity and long-term cardiovascular risk. Although high-sensitivity cardiac troponin (hs-cTn) is a marker of myocardial injury, its peripartum dynamics across HDP phenotypes are not well characterized.

**Methods:** A retrospective cohort study was conducted. Pregnant patients ≥18 years with samples within ±48 hours of delivery were included. Mixed-effects linear regression compared log10-transformed hs-cTnI levels and slopes across PEC, HTN, and normotensive controls (NC), adjusting for clinical covariates.

**Results:** Among 609 participants, median hs-cTnI increased significantly after delivery across all groups. Adjusted models showed higher hs-cTnI in PEC vs NC (β=0.266, p<0.001) and HTN (β=0.160, p=0.011), and in HTN vs NC (β=0.107, p=0.046). Slopes were steeper in PEC vs NC (β=0.005/h, p=0.013), but not between PEC and HTN.

**Conclusions:** These findings support the concept of delivery as a physiologic “stress test” and provide new insights into phenotype-specific cardiovascular burden in pregnancy.

## Introduction

Hypertensive disorders of pregnancy (HDP), including preeclampsia (PEC) and chronic/gestational hypertension (HTN), are major causes of fetal complications, maternal morbidity and predictors of future cardiovascular disease ^1^. High-sensitivity cardiac troponin (hs-cTn) is a marker of myocardial injury, and expert guidance cautions that elevations in pregnancy, peripartum, or in PEC should not be dismissed as physiologic ^2^. This guidance is supported by a large population (2358 patients) study showing no differences in hs-cTn between nonpregnant and pregnant women across trimesters ^3^, but without peripartum assessment, when cardiac burden peaks. Smaller peripartum studies used inconsistent phenotype stratification (separating PEC or pooling HDP) ^4–6^. To address this gap, we assessed hs-cTnI within a tight ±48h peripartum window, comparing PEC, HTN, and normotensive controls (NC).

## Methods

We conducted a retrospective cohort study at NewYork-Presbyterian/Weill Cornell Medical Center (IRB #1806019375). Participants were pregnant patients ≥18 years with leftover serum/EDTA plasma for routine clinical testing. Hs-cTnI was measured using Siemens Centaur (Limit of quantitation (LOQ): 2.5 ng/L; female 99^th^ percentile:37.0 ng/L). Exclusion criteria: postpartum PEC and prior cardiovascular diseases. Mixed-effects linear regression compared log10-transformed hs-cTnI levels and slopes across groups, adjusting for age, race, multifetal gestation, chronic hypertension, nulliparity, pregravid BMI, blood loss, and delivery type. Statistical analyses were performed in R version 4.2.1.

## Results

Among 609 participants (788 samples; 90 PEC/153 samples, 116 HTN/161 samples, and 403 NC/474 samples), baseline characteristics differed significantly. PEC patients were older, more often Black/African Americans, had higher rates of multifetal gestation, earlier delivery, more cesareans, greater blood loss, and lower infant birth weight than HTN and NC. Median hs-TnI were below LOQ before delivery but increased significantly afterward across all groups: 9.1 ng/L (IQR 4.8-26.6) for PE, 5.9 ng/L (IQR 2.6-24.0) for HTN, and 5.7 ng/L (IQR 2.5-13.8) for NC, respectively (p<0.001). Elevations above the female 99^th^ percentile occurred in 6.7% of PEC, 2.6% of HTN, and 1.5% of NC (p=0.02) (**Table 1**).

**Table 1.** Patient Demographic and Baseline Characteristics.

| Variable | All patients<br>(n=609) | PEC<br>(n=90) | HTN<br>(n=116) | NC<br>(n=403) | p value^ |
| --- | --- | --- | --- | --- | --- |
| Age, years, Median (IQR) | 34 (31-37) | 35 (32-38) | 34 (32-38) | 33 (31-36) | **0.007** |
| Multifetal, n (%) | 16 (2.6) | 8 (8.9) | 0 (0.0) | 8 (2.0) | **<0.001** |
| Chronic hypertension, n (%) | 29 (4.8) | 11 (12.2) | 18 (15.5) | 0 (0.0) | **<0.001** |
| Race, n (%) |  |  |  |  |  |
| *White* | 380 (62.4) | 45 (50.0) | 82 (70.7) | 253 (62.8) | **<0.001** |
| *Asian* | 81 (13.3) | 8 (8.9) | 13 (11.2) | 60 (14.9) |  |
| *Black or African American* | 34 (5.6) | 13 (14.4) | 8 (6.9) | 13 (3.2) |  |
| *Other Combinations not Described* | 65 (10.7) | 13 (14.4) | 6 (5.2) | 46 (11.4) |  |
| *American Indian or Alaska Nation* | 1 (0.2) | 0 (0.0) | 0 (0.0) | 1 (0.2) |  |
| *Declined* | 48 (7.9) | 11 (12.2) | 7 (6.0) | 30 (7.4) |  |
| Cesarean Section delivery, n (%) | 227 (37.3) | 54 (60.0) | 48 (41.4) | 125 (31.0) | **<0.001** |
| Blood loss volume, mL, Median (IQR) | 500 (206, 800) | 685 (424, 886) | 500 (293, 800) | 406 (200, 712) | **<0.001** |
| Baby weight, g, Median (IQR) | 3,235 (2,935, 3,570) | 2,730 (2,260, 3,125) | 3,220 (2,975, 3,580) | 3,303 (3,035, 3,628) | **<0.001** |
| Nulliparity, n (%) | 398 (65.4) | 66 (73.3) | 81 (69.8) | 251 (62.3) | 0.075 |
| Pregravid BMI, kg/ m<sup>2</sup>, Median (IQR) | 23.9 (21.4, 28.0) | 25.3 (22.5, 30.0) | 25.0 (21.8, 30.0) | 23.3 (21.1, 27.0) | **<0.001** |
| Delivery GA, weeks, Median (IQR) | 39.1 (38.0, 40.0) | 37.1 (35.7, 39.0) | 38.8 (37.7, 40.0) | 39.6 (38.7, 40.0) | **<0.001** |
| High-sensitivity troponin I level, ng/L |  |  |  |  |  |
| Before delivery, Median | 0 (0, 0) | 0 (0, 5.7) | 0 (0, 0) | 0 (0, 0) | **<0.001** |
| (IQR) |  |  |  |  |  |
| After delivery, Median (IQR) | 7.0 (3.4, 19.9) | 9.1 (4.8, 26.6) | 5.9 (2.6, 24.0) | 5.7 (2.5, 13.8) | 0.249 |
| Before vs. After (p value<sup>^</sup>) | **<0.001** | **<0.001** | **<0.001** | **<0.001** | - |
| Troponin I above 99<sup>th</sup> Percentile, n (%) | 15 (2.5) | 6 (6.7) | 3 (2.6) | 6 (1.5) | **0.020** |

Note: PEC: Preeclampsia; NC: Normal control; HTN: Chronic or gestational hypertension without PEC; IQR, Interquartile range; BMI, Body mass index; GA, Gestational age; “<sup>^</sup>”: Wilcoxon rank-sum test or Kruskal-Wallis test, Fisher’s exact test or Chi-square test as appropriate. Troponin I: Only used the highest value if a patient has multiple samples.

Mixed-effects analysis demonstrated higher log10-transformed hs-TnI levels in PEC compared with NC (β=0.318, 95%CI 0.228–0.408, p<0.001) and HTN (β=0.167, 95%CI 0.058–0.276, p=0.003), with HTN also higher than NC (β=0.151, 95%CI 0.061–0.240, p<0.001). Slopes were steeper in PEC (β=0.004/h, 95%CI 0.000–0.008, p=0.043) and HTN (β=0.004/h, 95%CI 0.000– 0.008, p=0.041) versus NC, but did not differ between PEC and HTN (p=0.968). After multivariable adjustment, PEC remained higher than NC (β=0.266, 95%CI 0.160–0.373, p<0.001) and HTN (β=0.160, 95%CI 0.036–0.283, p=0.011), and HTN remained higher than NC (β=0.107, 95%CI 0.002–0.212, p=0.046). Adjusted slope analysis confirmed a greater rise in PEC versus NC (β=0.005/h, 95%CI 0.001–0.009, p=0.013), with no slope differences between HTN and either group (**Figure 1**).

**Figure 1.**
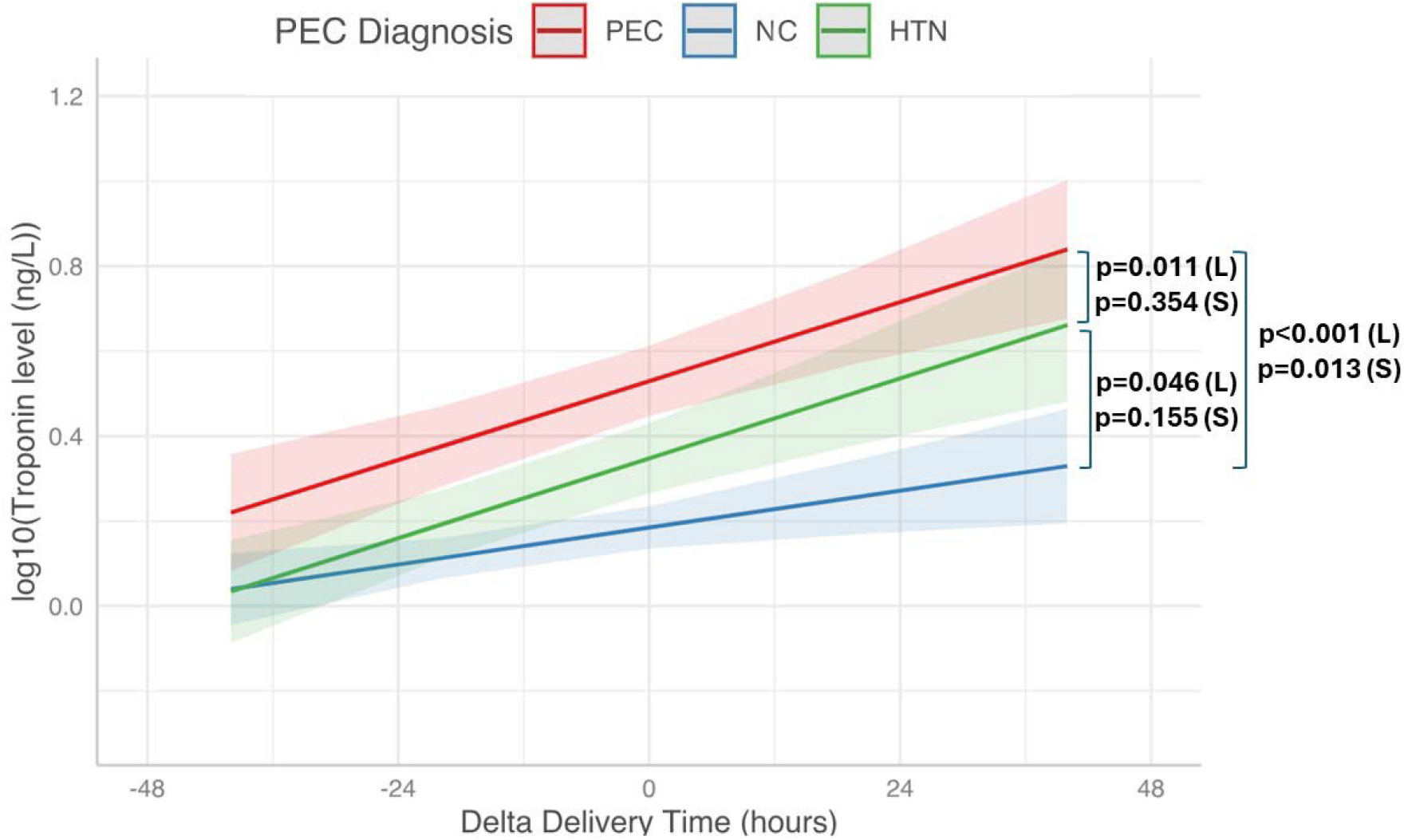
Log10-transformed hs-TnI trajectories within 48 hours of delivery in PEC (red), HTN (green), and NC (blue). Mixed-effects linear regression compared hs-TnI levels and slopes across groups (PEC: preeclampsia; HTN: chronic or gestational hypertension without PEC; NC: normotensive control). To enable log10 transformation, half of the minimum hs-TnI value was added to every value. Model estimates (solid lines, shaded 95% CI) are shown for 48 hours before and after delivery. Brackets on the right indicate comparisons of hs-TnI levels (L) and slopes (S) and p values are from multivariable-adjusted mixed-effects models. In multivariable models, PEC showed higher levels than both NC (p < 0.001) and HTN (p = 0.011), and HTN was higher than NC (p = 0.046). For slopes, PEC rose more steeply than NC (p = 0.013); HTN versus NC was significant only in univariate analysis (p = 0.041) but not after adjustment (p = 0.155), and no difference was observed between PEC and HTN (p = 0.354).

## Discussion

This study provides the first systematic assessment of hs-cTnI dynamics within the immediate peripartum window. PEC was associated with both higher levels and a steeper rise than NC, while HTN showed intermediate elevations. Increases above the 99^th^ % URL were rare despite increases in values pre to post delivery. These findings highlight peripartum as the point of maximal cardiovascular stress, when phenotype-specific strain emerges. The disproportionate hs-cTnI rise in PEC compared with HTN likely reflects its unique pathophysiology of endothelial dysfunction, anti-angiogenic imbalance, and microvascular injury, amplifying stress beyond elevated blood pressure alone. Our results complement prior epidemiologic evidence linking PEC to elevated long-term cardiovascular risk and delivery as a physiologic “stress test” for the maternal heart. These data also provide guidance for clinicians who care for patients during the delivery period. Limitations include the single-center design, limited longitudinal sampling, and lack of extended follow-up. Nonetheless, these findings fill an important gap and provide new insights into phenotype-specific maternal cardiac stress.

## Data Availability

All data produced in the present study are available upon reasonable request to the authors

## Author Contributions

*Concept and design*: Zhen Zhao and Allan S. Jaffe

*Acquisition, analysis, or interpretation of data*: Yaxin Li, Zhengming Chen, Allan S. Jaffe, Damien Gruson, Sarina He Yang, and Isla Racine

*Drafting of the manuscript*: Yaxin Li, Sarina He Yang, Isla Racine, Zhengming Chen, and Zhen Zhao

*Critical revision of the manuscript for important intellectual content*: Allan S. Jaffe, Damien Gruson

*Statistical analysis*: Zhengming Chen

*Obtained funding*: Zhen Zhao

*Administrative, technical, or material support*: Zhen Zhao

*Supervision*: Zhen Zhao

## References

1. Wu P, Haththotuwa R, Kwok CS, et al. Preeclampsia and Future Cardiovascular Health: A Systematic Review and Meta-Analysis. Circ Cardiovasc Qual Outcomes. Feb 2017;10(2)doi:10.1161/CIRCOUTCOMES.116.003497

2. Sarma AA, Aggarwal NR, Briller JE, et al. The Utilization and Interpretation of Cardiac Biomarkers During Pregnancy: JACC: Advances Expert Panel. JACC Adv. Aug 2022;1(3):100064. doi:10.1016/j.jacadv.2022.100064

3. Minhas AS, Echouffo-Tcheugui JB, Zhang S, et al. High-Sensitivity Troponin T and I Among Pregnant Women in the US-The National Health and Nutrition Examination Survey, 1999-2004. JAMA Cardiol. Apr 1 2023;8(4):406–408. doi:10.1001/jamacardio.2022.5601

4. Umazume T, Yamada S, Yamada T, et al. Association of peripartum troponin I levels with left ventricular relaxation in women with hypertensive disorders of pregnancy. Open Heart. 2018;5(2):e000829. doi:10.1136/openhrt-2018-000829

5. Kimura Y, Kato T, Miyata H, et al. Factors associated with increased levels of brain natriuretic peptide and cardiac troponin I during the peripartum period. PLoS One. 2019;14(2):e0211982. doi:10.1371/journal.pone.0211982

6. Morton A, Morton A. High sensitivity cardiac troponin I levels in preeclampsia.

